# Knowledge, attitude and preventive practices towards scabies infection among mothers of under-five children in Ibadan south-east local government area, oyo state, nigeria

**DOI:** 10.1101/2025.10.28.25339030

**Authors:** Melody Adefaka Abiodun, Mojisola M. Oluwasanu, Gabriel Ogunde

**Affiliations:** Department of Health Promotion and Education, Faculty of Public Health, College of Medicine, University of Ibadan, Ibadan, Nigeria; Department of Epidemiology and Medical Statistics, Faculty of Public Health, College of Medicine, University of Ibadan, Ibadan, Nigeria

## Abstract

Scabies, an infection caused by the infestation of the skin by *Sarcoptes scabiei* mites, which is endemic in low-resource settings, with under-five children severely affected. In Nigeria, factors such as overcrowding, poor sanitation, and limited healthcare access sustains the continued prevalence of the infection, contributing to significant morbidity and secondary bacterial infections. Despite its recognition as a neglected tropical disease by the World Health Organisation, gaps persist in understanding maternal knowledge, attitude, and preventive practices critical to mitigating transmission in semi-urban settings. This study investigated knowledge, attitude, and preventive practices toward scabies among mothers of under-five children in Ibadan South East, Local Government. A cross-sectional survey was conducted among 320 mothers of under-five children, selected via a three-stage sampling technique using a structured, interviewer-administered questionnaire. Results were presented using descriptive statistics and, multiple linear regression analysis at α^0.05^. Respondents’ mean age was 28.7±4.9. 48.4%, had good knowledge. A high proportion (76.5%) were unaware of skin-to-skin transmission. However, 79.3% recognised overcrowding as a risk factor. Attitude was predominantly negative (62.8%). Preventive practices were poor (83.1%); 42.5% shared cover clothes, and 24.4% avoided clinics despite detecting symptoms. Regression analysis showed that knowledge was significantly associated with age of the youngest child (β = –0.62 for age 2, *p* = 0.003; β = 0.54 for age 4, *p* = 0.017). Attitude was influenced by maternal age, ethnicity, child’s age, and income (*p* < 0.05). Preventive practices were significantly lower among single mothers (β = –1.22, *p* = 0.021). This study reveals the critical gaps in maternal knowledge, attitudes, and preventive practices toward scabies in urban Nigeria, driven by low education, economic constraints, and cultural practices. Misconceptions about transmission underscore the need for culturally tailored interventions. This can be addressed by implementing, community health education programmes, media campaigns and policy advocacy.

## Introduction

Scabies is an infection caused by the infestation of the skin by *Sarcoptes scabiei* mites, which is endemic in areas such as Egypt, Central America, South America, Africa, India, and South East Asia (Yirgu, Middleton, Tesfaye and Enbiale, 2023). This is commonly known as the seven-year itch, it is a contagious skin infection which can be transmitted from human beings to animals and vice-versa. The status of scabies as a neglected tropical disease was recognised by the World Health Organisation in 2017, which has increased research efforts and public health interventions (Engelman, Steer, McCarthy, Hay, Currie et al., 2019). The mite burrows into the host’s skin surface. This, in most cases, results in serious allergic inflammation. The present estimation is that upwards of 200 million people worldwide are affected at any given time (Chandler, David, Fuller, Claire, Engelman, Daniel, 2024). Whereas anybody, irrespective of age, sex, ethnicity, and socio-economic background, may be affected by the disease, it particularly causes illness among populations living in overcrowded conditions, especially in areas where access to healthcare is limited and in regions experiencing poverty (Romani, Lucia, Marks, Michael, Sokana, Oliver and Kakande, 2023).

The incubation period, from initial infection to development of symptoms, is approximately 2-6 weeks but may vary depending on the host’s immune status and the number of mites transferred (Mahe, Antoine, Hay, Roderick and Chosidow, 2023). The morbidity statistics of scabies peak among children, particularly those under-five children and of school age, and mainly in children from impoverished areas (Chandler, David, Fuller, 2019).

It is a common disease in Nigeria, mainly due to environmental factors like dense population, poor environmental sanitation, contaminated water, and soil (Ogunbiyi, Omigbodun, Owoaje, Odusan, Adekanle, 2022; Ayanlowo, Okesola, Kehinde, Adebola, Adebiyi, 2023). Olaniyi et al assessed the prevalence and associated risk factors for scabies infestation among children in some selected communities in Oyo State, Nigeria (Olaniyi, Adebowale, Oyewole, Adesina and Ogunkanmi, 2023). Overcrowding, low socio-economic status, poor personal hygiene and limited hygiene were some of the major risk factors for scabies infestation. The factors have sustained the continued prevalence of the infection, contributing to significant morbidity and secondary bacterial infections. Nweze, (2021) conducted a cross-sectional survey of 5,000 secondary school students drawn from 50 randomly selected schools across the three zones of Anambra State and reported an overall scabies prevalence of 10.0% (500/5,000); using a structured questionnaire and direct clinical examination the study identified sharing of beds, pillows and clothes, overcrowded sleeping arrangements and irregular bathing as important risk factors. These findings underline the need for targeted interventions, particularly those addressing these risk factors and improving access to effective treatment to control and prevent scabies within the region. Recent research has continued to identify the need to improve awareness and attitudes towards scabies among vulnerable populations. (Nweze, 2021). Scabies among under-five children is associated with intense itching, sleep disturbance, and skin sores, which can lead to secondary bacterial infections such as impetigo and cellulitis, sometimes progressing to kidney diseases. These effects compromise the well-being of young children, affect their growth and school readiness, and increase the burden on families. Though scabies generally do not pose a serious threat to life, early management and preventive measures are very significant in controlling their spread. Despite its recognition as a neglected tropical disease by the World Health Organisation, gaps persist in understanding maternal knowledge, attitude, and preventive practices critical to mitigating transmission in semi-urban settings.

Findings carried out in recent studies have shown that maternal knowledge was instrumental in the spread of the disease among under-five children. Engelman and Romani, 2024 identified that scabies predominantly affects under-five children, who face higher risks of secondary infections and developmental disturbances. Emanghe et al, (2024) also found that a significant relationship existed between the knowledge and practice of mothers and the burden of scabies among young children, which again makes maternal education and awareness very important in the control of scabies both at household and community levels.

The attitudes of mothers towards scabies prevention and management significantly influence infection rates among under-five children. In urban Nigerian settings, there is a gap in understanding mothers’ attitudes towards scabies prevention. This lack of information has limited the development of effective preventive strategies tailored to urban contexts. Ezugwu et al, (2021) highlighted this knowledge gap concerning local dynamics for scabies prevention in Nigeria, emphasising the need for more studies exploring urban settings and mothers’ attitudinal dispositions such as perceived severity of scabies, the stigma associated with skin diseases, trust in healthcare systems and self-efficacy in implementing preventive measures regarding the disease.

Preventable practices among mothers of under-five children play the most significant role in the transmission of the disease. These include poor personal hygiene practices, wrong and infrequent washing of clothes and bedding, and ignorance of the early signs and symptoms of the disease. According to Awogbenja et al, (2023), specific preventable practices in an urban setting in Nigeria, such as irregular use of soap for bathing, infrequent washing of shared beddings, reliance on traditional remedies over orthodox treatments, and delays in isolating infected family members, have not been reported, which is very necessary for the development of an appropriate intervention. Understanding these practices is essential for formulating pragmatic strategies for scabies prevention and control in urban Nigerian communities. This study investigated knowledge, attitude, and preventive practices toward scabies among mothers of under-five children in Ibadan South East, Local Government, Oyo State, Nigeria.

## Materials and Methods

### Study design

A cross-sectional study was conducted. This design was particularly suitable for this study as it allowed for the collection of data from the target population at a specific point in time (Setia, 2016). The main objective of the study was to explore the level of knowledge, attitude, and preventive practices of scabies infection among mothers of under-five children in Ibadan, South-East, Oyo State. This study design enabled the researcher to collect and analyse the data without omitting necessary details, offering a snapshot of the current state of knowledge, attitude, and preventive practices of scabies infection among the respondents.

### Study area and period

The study was conducted in Ibadan South East Local Government Area, a predominantly urban district within the larger Ibadan metropolis. Covering just 17 km², this LGA is characterized by very dense settlement patterns and evolving urban infrastructure. According to the most recent census, Ibadan South East is home to approximately 302,271 residents, making it one of the most congested areas in the city. Its compact geography and high population density present both significant challenges and opportunities for economic expansion, social development, and public health interventions. The study was conducted from December, 2024 to January, 2025.

### Study population, inclusion criteria

Mothers of under-five children residing in the selected communities within the Ibadan South East Local Government Area. All mothers of under-five children who consented to participate in the study were selected.

### Sample size, sampling technique

The sample size for this study was estimated from the Leslie Kish formula for a single proportion which is as follows; By considering 25% prevalence of knowledge about scabies infection, which was obtained from Olaniyi et al. (2023), margin of error (d) = 5%, and 5% significance level (α). The calculated minimum sample size was 288. After adding a 10% non-response rate, the final sample size was 320 participants.

### Data collection tools, and procedures

Data collection involved interviewer-administered questionnaires completed using KoboCollect, an electronic data collection tool. Before data collection, five (5) research assistants underwent comprehensive training on the proper administration of the questionnaire through KoboCollect. They were educated on respecting respondents’ rights and avoiding any violations. Additionally, they were instructed on how to interact with respondents, including obtaining informed consent and thoroughly explaining the study’s purpose to all stakeholders involved.

### Study variables

Key dependent variables were Knowledge, attitude, and preventive practices towards scabies infection.

Knowledge – Comprehensive understanding of scabies including its causes, how it spreads, common signs and symptoms, preventive strategies, and available treatments was assessed using a 9-item questionnaire. Participants scoring 0–5 were classified as having poor knowledge, while those scoring 6–9 were classified as having good knowledge. (Chandler and Fuller, 2019; Siddig and Hay, 2022).

Attitude – Participants’ perceptions and beliefs about scabies severity, their openness to taking preventive measures, willingness to seek medical care, sense of personal vulnerability, and views on community responsibility were measured with a 14-item Likert scale. Total scores of 0–8 indicated a negative attitude, whereas scores of 9–14 indicated a positive attitude. (Engelman et al., 2019).

Preventive Practices – Routine community behaviours to prevent scabies such as frequency of personal and household hygiene routines, use of protective measures for children, adherence to medical advice, and proactive steps during outbreaks were evaluated with a 13-item practice inventory. Participants scoring 0–7 were deemed to have poor preventive practices, and those scoring 8–13 were deemed to have good preventive practices (Dagne et al, 2023; CDC, 2023).

### Data analysis

Both descriptive and inferential statistical analyses were performed to test for differences in the Knowledge, Attitude, and Practice variables across socio-demographic categories, with each variable analysed separately. In addition, multiple linear regression models were estimated using the ‘regress’ command to explore relationships between the continuous outcome variable and relevant predictors. We analysed the data using STATA version 16.

### Ethics consideration

The research ethics application for conducting this study was reviewed and approved by the Oyo State Ministry of Health Research Ethics Committee with ethical approval no. (–Ref no. AD 13/479/555). The principles of beneficence and non-maleficence was equally observed throughout this study. Respondents were fully informed about the nature, benefits, and objectives of the study. Emphasis was given for voluntary participation, accompanied by the right to refuse or withdraw at any time without unfavourable consequences. Informed consents were obtained from all the respondents before interviews.

## Results

### 1. Socio-demographic characteristics

The study revealed the frequency distribution of the socio-demographic variables. A total of three hundred and twenty mothers of under-five children took part in this study. Table 1 shows the frequency distribution of the socio-demographic variables. Of the 320 participants, the average age (mean ± SD) was 28.7 ± 4.9 years, with 35.3% between 26-30 years old. Majority of the respondents (84.6%) were married while 10.6% were single. A large number of the respondents (60.9%) were Muslims while 39.0% were Christians. Few respondents 2.8% had no formal education, 8.4% had primary education, 58.4% had secondary education, and 30.3% had tertiary education. A little less than two third of the respondents 60.8% were self-employed, while 16.6% were Artisans. A large number of respondents 59.6% had children who were one year old, while 32.8% had children aged 2 years old. The mean age of the youngest child was 2.0 ± 1.1. Over half of the respondents’ 54.0% family income were less than ₦70000, and 15.9% were above ₦151,000.00.

**Table 1.** Respondents’ Socio-demographic Characteristics (N=320)

| **Variables** | **n** | **%** |
| --- | --- | --- |
| ***Average age (Mean ± SD)*** | ***28.7 ± 4.9*** |  |
| ***Age group (years)*** |  |  |
| $\leq 20$ | 13 | 4.1 |
| 21-25 | 72 | 22.5 |
| 26-30 | 113 | 35.3 |
| 31-35 | 103 | 32.1 |
| 36+ | 19 | 5.9 |
| ***Marital status*** |  |  |
| Divorced | 15 | 4.6 |
| Married | 271 | 84.6 |
| Single | 34 | 10.6 |
| ***Religion*** |  |  |
| Christianity | 125 | 39.0 |
| Islam | 195 | 60.9 |
| ***Ethnic Group*** |  |  |
| Yoruba | 278 | 86.8 |
| Non-Yoruba | 42 | 13.1 |
| ***Educational Status*** |  |  |
| No formal education | 9 | 2.8 |
| Primary | 27 | 8.4 |
| Secondary | 187 | 58.4 |
| Tertiary | 97 | 30.3 |
| ***Occupation*** |  |  |
| Student/unemployed | 31 | 9.7 |
| Self-employed | 194 | 60.8 |
| Artisan | 53 | 16.6 |
| Civil servant/Professional | 41 | 12.8 |
| ***Number of children under 5 years*** |  |  |
| 1 | 191 | 59.6 |
| 2 | 105 | 32.8 |
| 3 | 19 | 5.9 |
| 4 | 5 | 1.5 |
| ***Age of youngest child (Mean ± SD)*** | ***2.0 ± 1.1*** |  |
| ***Family size (total number of people living in your household)*** |  |  |
| 1-3 | 93 | 29.0 |
| 4-5 | 164 | 51.2 |
| ***Family income*** |  |  |
| < ₦70000 | 173 | 54.0 |
| ₦70k-150k | 96 | 30.0 |
| > ₦151k | 51 | 15.9 |

### 2. Knowledge of Scabies Infection

Knowledge of scabies infection among all study participants was measured on a 9-point scale with a mean score of 5.08 (SD = 1.46). Of the 320 study participants who responded to all items, 155 (48.4%) had good knowledge about scabies infection, and 165 (51.6%) had poor knowledge. Only 64 (20%) of respondents identified the cause of scabies and 75 (23.4%) described its mode of transmission. A large number of respondents 254 (79.3%) identified that people living in crowded conditions were at risk of contracting scabies. About two third of respondents 253 (70.0%) claimed that overcrowded living conditions encourage the spread of scabies infection.

**Table 2.** Knowledge of Scabies Infection (N=320)

| **Variables** | **Yes**<br>**n (%)** | **No**<br>**n (%)** |
| --- | --- | --- |
| Scabies is a contagious skin disease caused by Bacteria? | 64 (20%) | 256 (80%) |
| Scabies can be transmitted through skin to skin contact | 75 (23.4%) | 245 (76.6%) |
| Scabies infection can affect multiple family members simultaneously | 237 (74.1%) | 83 (25.9%) |
| People living in crowded conditions are at risk of contracting scabies | 254 (79.4%) | 66 (20.6%) |
| Overcrowded living conditions encourage the spread of scabies infection | 253 (79.1%) | 67 (20.9%) |
| Scabies symptoms appear within 2-6 weeks after infection | 72 (22.5%) | 248 (77.5%) |
| Intense itching, especially at night, is a common symptom of scabies | 113 (35.4%) | 207 (64.7%) |
| Secondary bacterial infections can result from scabies infection? | 198 (61.9%) | 122 (38.1%) |
| Benzyl benzoate lotion is the primary treatment for scabies infection | 188 (58.7%) | 132 (41.3%) |

### 3. Attitude towards Scabies Infection

Attitude of mothers of under-five children towards scabies infection was measured on a 14-point scale with a mean score of 7.75 (SD = 2.10) which two thirds (62.8%) had negative attitude and one third (37.2) had positive attitude. Majority (81.9%) agreed that scabies is a more serious health concern for children under-five children, and many (82.8%) agreed that scabies is easily prevented through good hygiene practices. 43.8% disagreed that scabies infectious was a sign of poor parenting. Almost half (44.7%) agreed that traditional treatments are more effective than orthodox treatments in treating scabies. 62.5% agreed that scabies infections can have long-term effects on a child’s health. About two fifths (42.7%) agreed that Scabies is less serious than other skin conditions that affect children and (40.6%) disagreed that that once a child has had scabies, they become immune to future infections.

**Table 3.** Attitude towards Scabies Infection.

| **Variables** | **Agree**<br>**n (%)** | **Undecided**<br>**n (%)** | **Disagree**<br>**n (%)** |
| --- | --- | --- | --- |
| Scabies is a more serious health concern for children under-five | 262 (81.8) | 14 (4.4) | 44 (13.8) |
| Scabies is easily preventable through good hygiene practices | 265 (82.8) | 23 (7.2) | 32 (10.0) |
| Scabies infections are a sign of poor parenting | 130 (40.6) | 50 (15.6) | 140 (43.8) |
| I feel comfortable discussing scabies with healthcare providers | 237 (74) | 37 (11.6) | 46 (14.4) |
| Traditional treatments are more effective than orthodox treatments in treating scabies | 143 (44.6) | 92 (28.8) | 85 (26.6) |
| Scabies is only a problem for people living in poverty | 112 (35) | 28 (8.8) | 180 (56.2) |
| All my family members would be treated if one person has scabies | 212 (66.2) | 12 (3.8) | 96 (30) |
| I would feel embarrassed if my child contracted scabies | 176 (55) | 21 (6.6) | 123 (38.4) |
| I feel confident in my ability to identify early signs of scabies in my child | 207 (64.9) | 50 (15.7) | 62 (19.4) |
| Seeking medical help for scabies is unnecessary | 41 (12.9) | 19 (5.9) | 259 (81.2) |

**Table 3. Attitude towards Scabies Infection (cont.)**
| **Variables** | **Agree**<br>**n (%)** | **Undecided**<br>**n (%)** | **Disagree**<br>**n (%)** |
| --- | --- | --- | --- |
| Scabies treatment is too expensive for most families in our community | 115 (35.9) | 87 (27.2) | 118 (36.9) |
| Scabies is less serious than other skin conditions that affect children | 135 (42.2) | 58 (18.1) | 127 (39.7) |
| Once a child has had scabies, they become immune to future infections | 119 (37.2) | 71 (22.2) | 130 (40.6) |

### 4. Preventive Practices against Scabies Infection

Preventive practices against scabies infection was measured on a 13-point item with mean score of 3.79 (SD = 1.72). Majority of respondents (83.1%) had poor practices while (16.9%) had good practices. Half of the respondents (57.5%) with children under-five children did not have scabies in the past, (42.5%) did have the infection. Two third (61.9%) bath their children daily, (23.1%) bath thrice daily, (14.1%) bath only once and a few (0.94%) bath their children multiple times. Below half (44.4%) change their children’s clothes twice daily, (29.4%) change thrice daily and (17.2%) do that multiple times daily. Half of the respondents (55%) wash their bed sheets weekly, one third (30.6%) wash every two weeks and a few (11.6%) wash daily. Half of respondents (59.1%) wash their body towels weekly, (25.6%) wash daily and (11.3%) was every two weeks. A few (5.6%) do share underwear, while (42.5%) share cover clothes. Two third of the respondents (75.6%) visits the clinics when they notice any difference on their children’s skin while (24.4%) do not.

**Table 4.** Preventive Practices against Scabies Infection (N=320)

| **Variables** | **n** | **%** |
| --- | --- | --- |
| Has any of your children under-five had scabies in the past |  |  |
| *No* | 184 | 57.5 |
| *Yes* | 136 | 42.5 |
| Do you regularly inspect your children's skin for signs of rashes or itching |  |  |
| *No* | 59 | 18.4 |
| *Yes* | 261 | 81.6 |
| How many times do you bathe your children daily |  |  |
| *Multiple times* | 3 | 0.94 |
| *Once* | 45 | 14.1 |
| *Thrice* | 74 | 23.1 |
| *Twice* | 198 | 61.9 |
| How often do you change your children's clothes daily |  |  |
| *Multiple times* | 55 | 17.2 |
| *Once* | 29 | 9.1 |
| *Thrice* | 94 | 29.4 |
| *Twice* | 142 | 44.4 |
| How often do you wash bed sheets |  |  |
| *Daily* | 37 | 11.6 |
| *Every two weeks* | 98 | 30.6 |
| *More than two weeks* | 9 | 2.8 |
| *Weekly* | 176 | 55.1 |
| How often do you wash body towels |  |  |
| *Daily* | 82 | 25.6 |
| *Every two weeks* | 36 | 11.3 |
| *More than two weeks* | 13 | 4.1 |
| *Weekly* | 189 | 59.1 |
| Do you spread out towels every time after use |  |  |
| *No* | 72 | 22.5 |
| *Yes* | 248 | 77.5 |
| How often do you change your pillowcases |  |  |
| *Daily* | 48 | 15.1 |
| *Every two weeks* | 88 | 27.5 |
| *More than two weeks* | 18 | 5.6 |
| *Weekly* | 166 | 51.9 |

**Table 4. Preventive Practices against Scabies Infection (cont.)**
| **Variables** | **Frequency** | **Percentage** |
| --- | --- | --- |
| How often do you spread out your mattress/sleeping mat |  |  |
| *Daily* | 72 | 22.5 |
| *Every two weeks* | 49 | 15.3 |
| *More than two weeks* | 97 | 30.3 |
| *Weekly* | 102 | 31.9 |
| Do you share a bathing sponge |  |  |
| *No* | 217 | 67.8 |
| *Yes* | 103 | 32.2 |
| Do your child(ren) share underwear |  |  |
| *No* | 302 | 94.4 |
| *Yes* | 18 | 5.6 |
| Do you share cover clothes |  |  |
| *No* | 184 | 57.5 |
| *Yes* | 136 | 42.5 |
| Do you visit clinics when you notice any difference on your child(ren) skin |  |  |
| *No* | 78 | 24.4 |
| *Yes* | 242 | 75.6 |

### 5. Simple Linear Regression Model of Factors that are associated with Knowledge of Scabies Infection among Mothers of Under-five children

A simple linear regression analysis was conducted to identify predictors of good knowledge of scabies infection among mothers of under-five children (Table 5). The model showed that most socio-demographic variables were not significantly associated with knowledge levels (*p* > 0.05). However, two variables emerged as significant predictors. Mothers whose youngest child was two years old had significantly lower knowledge scores (β = –0.62, *p* = 0.003), while those with four-year-olds had significantly higher scores (β = 0.54, *p* = 0.017). As these variables yielded *p* < 0.05, we reject the null hypothesis.

**Table 5.** Simple Linear Regression Model of Factors that are associated with Good Knowledge of Scabies Infection among Mothers of Under-five children.

| **Variables** | **Coef.** | **Std. Err.** | **95% CI** | **p-value** |
| --- | --- | --- | --- | --- |
| ***Age group (years)*** |  |  |  |  |
| ≤ 20 | 0.00 | - | - | - |
| 21-25 | -0.54 | 0.43 | -0.91-0.80 | 0.917 |
| 26-30 | 0.24 | 0.42 | -0.05-1.08 | 0.570 |
| 31-35 | 0.38 | 0.42 | -0.45-1.22 | 0.367 |
| 36+ | 0.52 | 0.52 | -0.50-1.55 | 0.320 |
| ***Marital status*** |  |  |  |  |
| Divorced | 0.00 | - | - | - |
| Married | -0.55 | 0.38 | -1.32-0.20 | 0.149 |
| Single | -0.48 | 0.45 | -1.37-0.40 | 0.287 |
| ***Religion*** |  |  |  |  |
| Christianity | 0.00 | - | - | - |
| Islam | -0.24 | 0.16 | -0.57-0.08 | 0.144 |
| ***Ethnic Group*** |  |  |  |  |
| Yoruba | 0.00 | - | - | - |
| Non-Yoruba | 0.32 | 0.24 | -0.14-0.79 | 0.179 |
| ***Educational Status*** |  |  |  |  |
| No formal education | 0.00 | - | - | - |
| Primary | 0.66 | 0.56 | -0.43-1.77 | 0.236 |
| Secondary | 0.72 | 0.49 | -0.25-1.6 | 0.149 |
| Tertiary | 0.87 | 0.50 | -0.12-1.8 | 0.087 |
| ***Occupation*** |  |  |  |  |
| Student/unemployed | 0.00 | - | - | - |
| Self-employed | 0.00 | 0.27 | -0.54-0.55 | 0.975 |
| Artisan | -0.57 | 0.32 | -1.21-0.66 | 0.079 |
| Civil servant/Professional | 0.08 | 0.34 | -0.59-0.75 | 0.810 |
| ***Age of youngest child*** |  |  |  |  |
| 1 | 0.00 | - | - | - |
| 2 | -0.62 | 0.20 | -1.04, -0.21 | **0.003** |
| 3 | 0.10 | 0.23 | -0.35-0.55 | 0.664 |
| 4 | 0.54 | 0.22 | 0.09-0.98 | **0.017** |
| ***Family size*** |  |  |  |  |
| 1-3 | 0.00 | - | - | - |
| 4-5 | -0.19 | 0.18 | -0.56-0.17 | 0.304 |
| 6 or more | -0.03 | 0.23 | -0.50-0.42 | 0.867 |
| ***Family income*** |  |  |  |  |
| < ₦70000 | 0.00 | - | - | - |
| ₦70k-150k | 0.22 | 0.18 | -0.14-0.58 | 0.232 |
| > ₦151k | -0.05 | 0.23 | -0.51-0.58 | 0.808 |

### 6. Simple Linear Regression Model of Factors that are associated with Adequate Attitude towards Scabies Infection among Mothers of Under-five children

A simple linear regression analysis (Table 6) showed that several variables were significant predictors of mothers’ attitude scores. Compared to mothers aged 20 years, those aged 21–25 (β = –1.56, *p* = 0.014) and 31–35 (β = –1.27, *p* = 0.040) had significantly lower attitude scores. Non-Yoruba mothers reported more positive attitudes than Yoruba mothers (β = 0.69, *p* = 0.047). Regarding the age of the youngest child, mothers of two-year-olds (β = –0.81, *p* = 0.008) and three-year-olds (β = –0.95, *p* = 0.005) had lower attitude scores than those of one-year-olds. Finally, higher household income was associated with better attitudes: ₦70 000–150 000 (β = 0.70, *p* = 0.008) and >₦150 000 (β = 1.10, *p* = 0.001) compared to <₦70 000. As all these predictors had *p* < 0.05, we reject the null hypothesis.

**Table 6.** Simple Linear Regression Model of Factors that are associated with Adequate Attitude towards Scabies Infection among Mothers of Under-five children.

| **Variables** | **Coef.** | **Std. Err.** | **95% CI** | **p-value** |
| --- | --- | --- | --- | --- |
| ***Age group (years)*** |  |  |  |  |
| ≤ 20 | 0.00 | - | - | - |
| 21-25 | -1.56 | 0.63 | -2.80, -0.31 | **0.014** |
| 26-30 | -0.96 | 0.61 | -2.17-0.24 | 0.116 |
| 31-35 | -1.27 | 0.61 | -2.48, -0.05 | **0.040** |
| 36+ | -1.08 | 0.75 | -2.56-0.40 | 0.153 |
| ***Marital status*** |  |  |  |  |
| Divorced | 0.00 | - | - | - |
| Married | 0.19 | 0.56 | -0.90-1.29 | 0.730 |
| Single | 0.55 | 0.65 | -0.73-1.84 | 0.397 |
| ***Religion*** |  |  |  |  |
| Christianity | 0.00 | - | - | - |
| Islam | -0.37 | 0.24 | -0.84-0.10 | 0.122 |
| ***Ethnic Group*** |  |  |  |  |
| Yoruba | 0.00 | - | - | - |
| Non-Yoruba | 0.69 | 0.34 | 0.00-1.37 | **0.047** |
| ***Educational Status*** |  |  |  |  |
| No formal education | 0.00 | - | - | - |
| Primary | -0.62 | 0.81 | -2.22-0.96 | 0.438 |
| Secondary | -0.23 | 0.71 | -1.64-1.18 | 0.748 |
| Tertiary | 0.18 | 0.73 | -1.26-1.62 | 0.803 |
| ***Occupation*** |  |  |  |  |
| Student/unemployed | 0.00 | - | - | - |
| Self-employed | -0.36 | 0.40 | -1.16-0.42 | 0.360 |
| Artisan | -0.32 | 0.47 | -1.25-0.60 | 0.490 |
| Civil servant/Professional | 0.61 | 0.49 | -0.35-1.59 | 0.214 |
| ***Age of youngest child*** |  |  |  |  |
| 1 | 0.00 | - | - | - |
| 2 | -0.81 | 0.30 | -1.42, -0.21 | **0.008** |
| 3 | -0.95 | 0.33 | -1.62, -0.29 | **0.005** |
| 4 | -0.38 | 0.33 | -1.03-0.26 | 0.248 |
| ***Family size*** |  |  |  |  |
| 1-3 | 0.00 | - | - | - |
| 4-5 | 0.11 | 0.27 | -0.42-0.65 | 0.667 |
| 6 or more | -0.45 | 0.34 | -1.12-0.22 | 0.189 |
| ***Family income*** |  |  |  |  |
| < ₦70000 | 0.00 | - | - | - |
| ₦70k-150k | 0.70 | 0.26 | 0.18-1.22 | **0.008** |
| > ₦151k | 1.10 | 0.32 | 0.45-1.74 | **0.001** |

### 7. Simple Linear Regression Model of Factors that are associated with Preventive Practices of Scabies Infection among Mothers of Under-five children

A Simple linear regression model (Table 7) showed that four factors significantly predicted mothers’ preventive practice scores. Compared with divorced mothers, single mothers had significantly lower scores (β = –1.22, p = 0.021). Mothers with tertiary education also scored lower (β = –0.45, p = 0.044). Regarding the age of the youngest child, those with three-year-olds scored higher (β = 0.75, p = 0.007), whereas mothers of four-year-olds scored lower (β = –0.54, p = 0.045). Finally, in the middle income bracket (₦70 000–150 000) was associated with higher preventive practice scores (β = 0.43, p = 0.047). As each of these predictors had p < 0.05, we reject the null hypothesis.

**Table 7.** Simple Linear Regression Model of Factors that are associated with Preventive Practices of Scabies Infection among Mothers of Under-five children.

| **Variables** | **Coef.** | **Std. Err.** | **95% CI** | **p-value** |
| --- | --- | --- | --- | --- |
| ***Age group (years)*** |  |  |  |  |
| ≤ 20 | 0.00 | - | - | - |
| 21-25 | -0.56 | 0.51 | -1.58-0.44 | 0.272 |
| 26-30 | 0.04 | 0.50 | -0.94-1.03 | 0.930 |
| 31-35 | -0.18 | 0.50 | -1.17-0.81 | 0.716 |
| 36+ | -0.57 | 0.61 | -1.79-0.63 | 0.350 |
| ***Marital status*** |  |  |  |  |
| Divorced | 0.00 | - | - | - |
| Married | -0.32 | 0.45 | -1.21-0.56 | 0.473 |
| Single | -1.22 | 0.52 | -2.27, -0.18 | **0.021** |
| ***Religion*** |  |  |  |  |
| Christianity | 0.00 | - | - | - |
| Islam | -0.15 | 0.19 | -0.54-0.23 | 0.435 |
| ***Ethnic Group*** |  |  |  |  |
| Yoruba | 0.00 | - | - | - |
| Non-Yoruba | 0.23 | 0.28 | -0.32-0.79 | 0.407 |
| ***Educational Status*** |  |  |  |  |
| No formal education | 0.00 | - | - | - |
| Primary | 0.18 | 0.66 | -1.11-1.48 | 0.780 |
| Secondary | -0.52 | 0.58 | -1.67-0.63 | 0.375 |
| Tertiary | -0.45 | 0.59 | -1.63-0.72 | **0.044** |
| ***Occupation*** |  |  |  |  |
| Student/unemployed | 0.00 | - | - | - |
| Self-employed | 0.46 | 0.33 | -0.19-1.12 | 0.164 |
| Artisan | 0.73 | 0.39 | -0.33-1.50 | 0.061 |
| Civil servant/Professional | 0.48 | 0.41 | -0.32-1.29 | 0.241 |
| ***Age of youngest child*** |  |  |  |  |
| 1 | 0.00 | - | - | - |
| 2 | -0.20 | 0.24 | -0.69-0.28 | 0.419 |
| 3 | 0.75 | 0.27 | 0.20-1.29 | **0.007** |
| 4 | -0.54 | 0.27 | -1.03-0.26 | **0.045** |
| ***Family size*** |  |  |  |  |
| 1-3 | 0.00 | - | - | - |
| 4-5 | -0.06 | 0.22 | -0.50-0.37 | 0.771 |
| 6 or more | 0.43 | 0.28 | -0.11-0.98 | 0.124 |
| ***Family income*** |  |  |  |  |
| < ₦70000 | 0.00 | - | - | - |
| ₦70k-150k | 0.43 | 0.21 | 0.00-0.86 | **0.047** |
| > ₦151k | 0.09 | 0.27 | -0.44-0.63 | 0.722 |

## Discussions

This study examined the knowledge, attitudes, and preventive practices of mothers of under-five children toward scabies infection in Ibadan South-East, Oyo State, Nigeria. The findings reveal substantial gaps in knowledge, attitude, and preventive practices, all of which reinforce the ongoing burden of scabies in low-resource settings. Knowledge of scabies was notably poor, with less than one-third of respondents identifying the mite Sarcoptes scabiei as the causative agent, and only a quarter recognizing prolonged skin-to-skin contact as the major route of transmission. Instead, more than half attributed scabies to bacterial causes or environmental conditions, echoing findings from southern Nigeria where cultural and environmental explanations predominated (Emanghe et al., 2024). The recognition of overcrowding as a risk factor by two-thirds of respondents indicates partial awareness of epidemiologic drivers, yet persistent misconceptions underscore weaknesses in health education and entrenched cultural beliefs linking skin diseases to supernatural causes. Evidence from community-based interventions in Ethiopia and Ghana demonstrates that context-specific and culturally sensitive education can significantly shift perceptions, suggesting that similar models could be adapted locally (Tadesse et al., 2021; Adekeye et al., 2020).

Attitudes toward scabies further compound these challenges. Nearly two-thirds of respondents expressed negative perceptions, often shaped by stigma and the belief in the superiority of traditional remedies. Fear of social exclusion discourages disclosure and care-seeking, reflecting patterns reported in Ghana where stigma impedes timely treatment (Amoako et al., 2024). In contrast, studies in Kenya show that mothers engaged in sustained peer-led outreach are more likely to hold positive views and seek evidence-based treatment (Mwangi et al., 2024). These results highlight the importance of leveraging trusted local actors including religious leaders and traditional healers in order to counter stigma, reinforce accurate explanations, and promote trust in health services.

Although hygiene is generally considered important, preventive practices remain inconsistent. Weekly washing of bedding is reported by just over half of mothers, while clothing and towel sharing within households is still widespread. Such practices perpetuate mite transmission and align with previous reports from Ethiopia where limited sanitation constrained scabies control (Ayele et al., 2023). Economic and infrastructural barriers such as irregular water supply and limited access to laundry materials are widely cited, reflecting the structural dimensions of disease prevention. Integrated approaches that strengthen water, sanitation, and hygiene (WASH) infrastructure alongside community education have been shown to substantially reduce scabies prevalence in children (Azene et al., 2020). These findings underscore that individual-level behaviour change will remain fragile unless supported by broader system-level improvements.

## Conclusion

Scabies remains an important but under-recognized public health concern among under-five children in Ibadan South-East. This study shows that poor knowledge, negative attitudes, inconsistent preventive practices collectively sustain transmission. Addressing these gaps requires integrated strategies that combine culturally tailored health education, stigma reduction, and strengthened WASH infrastructure. Engaging trusted community actors such as religious leaders and traditional healers, alongside health professionals, fosters trust and improves care-seeking. Implementing multi-platform communication and system-level support is essential to reduce the burden of scabies in children and to inform broader control strategies across similar low-resource settings in sub-Saharan Africa.

## Supporting information

**S1 Questionnaire. Knowledge, attitude and preventive practices towards scabies infection among mothers of under-five children** (DOCX)

## Data Availability

All de-identified participant-level data necessary to reproduce the results reported in this manuscript are provided as Supporting Information with this submission (S1_Data.xlsx).

## Acknowledgements

The authors thank all the participants who took part in this study voluntarily and provided their information.

## Author Contributions

**Conceptualization:** Melody Adefaka Abiodun, Mojisola M. Oluwasanu

**Data curation:** Melody Adefaka Abiodun, Mojisola M. Oluwasanu

**Formal analysis:** Gabriel Ogunde, Melody Adefaka Abiodun, Mojisola M. Oluwasanu

**Methodology:** Melody Adefaka Abiodun, Mojisola M. Oluwasanu

**Project administration:** Melody Adefaka Abiodun, Mojisola M. Oluwasanu

**Supervision:** Mojisola M. Oluwasanu

**Writing – original draft:** Melody Adefaka Abiodun, Mojisola M. Oluwasanu **Writing – review & editing:** Melody Adefaka, Mojisola M. Oluwasanu

